# Heterogeneous immunological recovery trajectories revealed in post-acute COVID-19

**DOI:** 10.1101/2021.03.19.21254004

**Authors:** Yapeng Su, Dan Yuan, Daniel G. Chen, Kai Wang, Jongchan Choi, Chengzhen L. Dai, Sunga Hong, Rongyu Zhang, Jingyi Xie, Sarah Li, Kelsey Scherler, Ana Jimena Pavlovitch-Bedzyk, Shen Dong, Christopher Lausted, Rachel H. Ng, Inyoul Lee, Shannon Fallen, Sergey A. Kornilov, Priyanka Baloni, Venkata R. Duvvuri, Kristin G. Anderson, Jing Li, Fan Yang, Clifford Rostomily, Pamela Troisch, Brett Smith, Jing Zhou, Sean Mackay, Kim Murray, Rick Edmark, Lesley Jones, Yong Zhou, Lee Rowen, Rachel Liu, William Chour, William R. Berrington, Julie A Wallick, Heather A Algren, the ISB-Swedish COVID19 Biobanking Unit, Terri Wrin, Christos J. Petropoulos, Wei Wei, Nathan D. Price, Naeha Subramanian, Jennifer Hadlock, Andrew T. Magis, Antoni Ribas, Lewis L. Lanier, Scott D. Boyd, Jeffrey A. Bluestone, Leroy Hood, Raphael Gottardo, Philip D. Greenberg, Mark M. Davis, Jason D. Goldman, James R. Heath

**Author notes:** These authors contributed equally to this work (Yapeng Su, Dan Yuan, Daniel G. Chen).

## Abstract

The immunological picture of how different patients recover from COVID-19, and how those recovery trajectories are influenced by infection severity, remain unclear. We investigated 140 COVID-19 patients from diagnosis to convalescence using clinical data, viral load assessments, and multi-omic analyses of blood plasma and circulating immune cells. Immune-phenotype dynamics resolved four recovery trajectories. One trajectory signals a return to pre-infection healthy baseline, while the other three are characterized by differing fractions of persistent cytotoxic and proliferative T cells, distinct B cell maturation processes, and memory-like innate immunity. We resolve a small panel of plasma proteins that, when measured at diagnosis, can predict patient survival and recovery-trajectory commitment. Our study offers novel insights into post-acute immunological outcomes of COVID-19 that likely influence long-term adverse sequelae.

## Introduction

While the COVID-19 pandemic has resulted in over 2.3 million deaths worldwide, approximately 98% of infected individuals survive the infection. However, many survived patients either do not fully recover or only slowly regain their pre-infection baseline health status (1). Post-acute COVID-19 defines patients with symptoms extending beyond three weeks from initial onset, and chronic as beyond 12 weeks (2, 3). Although acute disease has been deeply characterized in several studies (4–10), most studies on convalescent patients have been centered on development and duration of immunological memory (11–14), less attention has been paid to capturing more comprehensive immunological views of the diverse recovery trajectories experienced by COVID-19 patients. However, such thorough longitudinal immunophenotyping may provide key insights for understanding long-term effects of COVID-19.

We investigated the recovery of 140 COVID-19 patients representing the full range of infection severities, through deep, multi-omic analyses of serial blood draws compared against healthy controls (Fig. 1A, table S1). Blood samples were collected at clinical diagnosis (T1, diagnosis), around 1 week later during acute disease (T2, acute), and 2-3 months later (T3, convalescent) when almost all patients had recovered from symptoms of acute disease. Each blood draw is analyzed for signatures of protective antibody immunity, large panels of plasma proteins and metabolites, and single-cell multi-omic analyses of peripheral blood mononuclear cells (PBMCs). Each blood draw was also paired with nasal swab measures of viral load and plasma viral RNAemia. These multi-omic and functional datasets were integrated within the context of electronic health records (EHR) of the same patients to connect clinical history with immunological recovery outcomes (Fig. 1A). We were able to stratify each studied patient into one of four COVID-19 recovery trajectories. Only one trajectory describes a return to a pre-infection, healthy baseline by T3.

**Fig. 1.**
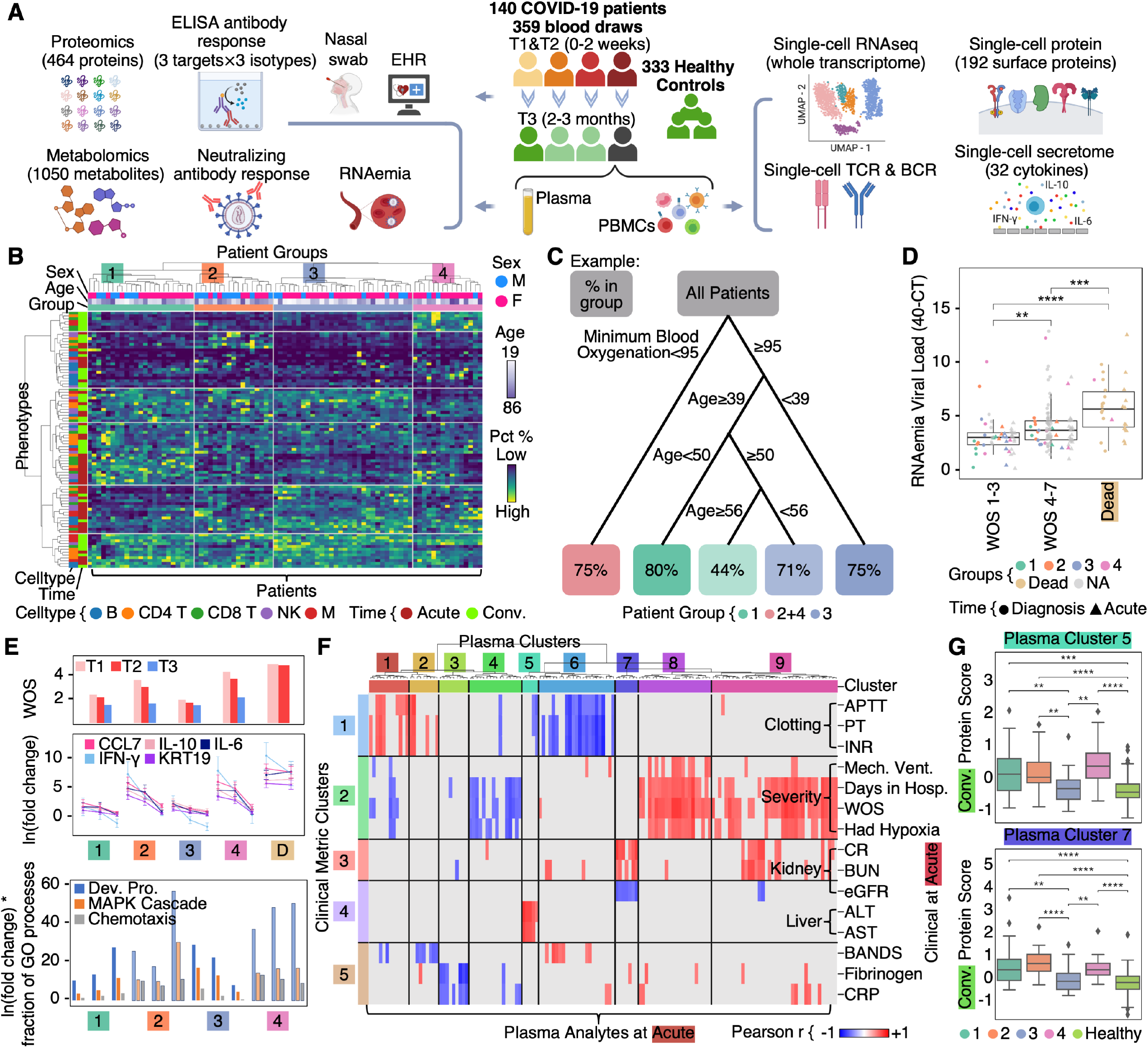
Overview of the Longitudinal Multi-Omic Analysis of Immune Responses in COVID-19 Patients. (A) Overview of ISB/Swedish INCOV Study design. The various analytic assays run on plasma and isolated PBMCs are shown. Detailed patient information listed in table S1. (B) Hierarchical clustering of patients based on immune cell subtypes at T2 and T3 resolved four patient-group trajectories. Each column (row) corresponds to the percentage of an immune subtype (patient) at acute (T2) or convalescence (T3). Top three rows indicate patient sex, age and patient group. Left two columns indicate major immune class and measurement time. Heatmap keys are provided beneath or to the right of the heatmap Full list of subpopulations and their percentages provided in table S2. (C) Decision tree classification model showing the top variables that discriminate patient groups using demographic and clinical variables recorded at T1 and T2. Colors of boxes the majority patient group (see color key) for the Leaf-node. Percentages within boxes indicate prediction accuracy of patient groups for that leaf node. Patient numbers per leaf node are listed in table S3. (D) RNAemia viral load analysis. Boxplot showing quantified plasma viral loads for samples of different severities at diagnosis (T1) and acute (T2). Y-axis unit is 40-CT (cycle threshold). Data points were color coded by patient groups. Data points from patients with no group assignments due to unavailable T3 blood draws are shown in grey. This data is listed in table S4. (E) Plasma proteomic analysis across patient groups. Top (bar plot), disease severity of each group per time point, plotted as mean±SEM. Middle (line plot), plasma abundance of five severity-related proteins (quantified by Z-scores, Supplemental Methods) per patient group. From left to right, each group of lines shows data at T1, T2, and T3, plotted as mean±SEM. Bottom, gene ontology (GO) score (Supplemental Methods) of three biological processes (infection responses) in each patient group. The three groups of bars for each patient group represent data at T1, T2, and T3. (F) Inter-omic correlations between plasma analytes and clinical data. Each row is a selected clinical measurement from electronic health records and each column is a plasma protein or metabolite. Top correlative clinical metrics are shown, full map shown in Fig. S1D. Color represents Pearson correlation coefficient of row and column variables (see color key at bottom). Significant correlations (|r|≥0.5 and p-value<0.05, Supplemental Methods) are shown. (G) Levels of plasma proteins in cluster 5 and 7 across patient groups at T3. Boxplot center, median; box limits, 25th and 75th percentiles; whiskers, 1.5× interquartile range (IQR). *p-value<0.05, **p-value<0.01, ***p-value<0.001, ****p-value<0.0001.

## Results

### Peripheral immune cell composition dynamics reveal four distinct recovery trajectories

We used single-cell transcriptomes of PBMCs collected from all patients (all timepoints) to identify major immune cell classes; visualized as a two-dimensional projection via uniform manifold approximation and projection (UMAP) (15) (Fig. S1A). Each class was further classified into subtypes based on global transcriptomic profiles (Fig. S1A). Immune cell compositions widely varied across patients within and between time points (table S2). We utilized these composition dynamics to classify patients into four immunological trajectory groups (Fig. 1B). Notably, while older patients are at higher risk of fatal COVID-19 (both in general (16) and in this cohort), age differences were only significant between groups 2 and 3 (Fig. S1B). The groupings independently reflected COVID-19 infection severity. Groups 2 and 4 were enriched for more severe COVID-19 patients, as defined by the WHO Ordinal Scale Score (WOS) (17), relative to groups 1 and 3 (Fig. S1B). This underscores disease severity as a major contributor to a patient’s immunological outcome at convalescence, although other factors exist.

Recursive partitioning of data from patient EHR (co-morbidities, medications, infection severity, etc.) was analyzed for clinical determinants of the immunological groupings (Fig. 1C, Supplemental Methods). These clinial metrics did not resolve differences between patient groups 2 and 4. The main differentiator was low levels of minimum blood oxygen (<95) were observed in groups 2 and 4. Age was the one variable that further resolved patient groupings. This analysis suggests clinical metrics track with immunological groupings.

Additionally, post-COVID-19 healthcare encounters for groups 2 and 4 were notable for respiratory symptoms and signs including fatigue, dyspnea, dyspnea on exertion, nausea/vomiting, the need for supplemental oxygen and/or ongoing mechanical ventilation (47% in Group 2 and 40% in Group 4). However, observations were only available for patients who had additional healthcare visits between T2 and T3. Additional research is needed to characterize patterns of longitudinal sequelae, compare results with SARS-CoV-2 negative reference populations, and investigate underlying etiology.

Viral load from qPCR analysis of nasal swabs correlates with COVID-19 infection severity (18) and mortality (19). RNAemia (detection of viral-RNA in plasma) also correlates with infection severity (20), but is only detected in a subset of patients. RNAemia assays discriminated between fatal (D), severe (WOS=4-7) and mild (WOS=1-3) disease at T1 and T2 (Fig. 1D).

### Plasma multi-omics integrated with clinical data reveal divergent outcomes across patient groups

Figure. 1E displays kinetic changes, per patient group, in five proteins reported to increase with acute infection severity (4, 9). Here, the y-axis represents natural log-fold changes above or below the value (set to 0) expected for matched non-COVID-19 healthy controls (Supplemental Methods). Notably, all groups show decreasing trends implying COVID-19 recovery (Fig. 1E top and middle). In contrast, many other measured proteins behave otherwise (table S5). We grouped proteins into specific gene ontology (GO) processes shared across all groups (Supplemental Methods) collectively representing ≥200 proteins. “Developmental Processes”, for example, reflects construction of an immune response during acute disease, and post-infection tissue repair. For the two mildly infected groups, group 3 exhibited an infection response at T1 that returned to near-healthy by T3, while group 1 exhibited the opposite trend (Fig. 1E bottom). Groups 2 and 4 exhibited strong responses at T1 that remained high at T3 (Fig. 1E bottom). This comprehensive proteomic view suggests continued engagement of immune processes in groups 1, 2 and 4.

Significant metabolic changes are also observed across patient groups at T3, including elevated pro-inflammatory metabolites (21, 22) in group 4, and immunosuppressive metabolites in group 2 (23–26) (Fig. S1C, table S6).

We explored for relationships between plasma analytes (proteins and metabolites) with clinical measures at T2 (Supplemental Methods) to identify surrogate markers for clinical metrics, which are typically less available at T3. Unsupervised clustering of correlation patterns grouped clinical measures via function (e.g. clotting metrics-row-cluster 1) (row clusters of Fig. 1F, Fig. S1D) and resolved correlated clusters of plasma analytes (columns of Fig. 1F, Fig. S1D, table S7). For example, plasma cluster 4 negatively correlates with clinical-severity-metrics, while clusters 8 and 9, which include IL-6 and TNF (table S7), positive-correlated with severity. Clusters 5 and 7 correlated with clinical metrics of liver (ALT, AST) and kidney (creatinine, BUN) damage, respectively (Fig. 1F), and were elevated in patient groups 1, 2 and 4 at T3 (Fig. 1G). These elevations suggest possible continued kidney and liver damage in some patients at T3, and again imply a slow recovery for certain patients.

### Patients with history of severe COVID-19 can have cytotoxic and proliferative T cells in convalescence

We analyzed for CD8+ and CD4+ T cell phenotype differences across the patient groups. We projected CD8+ and CD4+ T cell sc-CITE-seq data onto separate UMAP representations to identify distinct T cell phenotypes (Fig. 2A, Fig. S2A-C, S3A-C). Clonal expansion in group 2 was high for CD8+ and CD4+ T cells at T2 and T3 (Fig. S2D, S3D). Two T cell phenotypes previously reported to positively-correlate with disease severity(4, 8) remained unexpectedly enriched in some convalescent patients (Fig. 2B, Fig. S2A-C, E, S3A-C, E, F). The first are cytotoxic CD8+ and CD4+ T cells, both with upregulated CD45RA protein and high clonal expansion levels. The second is a hybrid (proliferative-exhausted) phenotype with elevated levels of proliferative (MKI67) and exhaustion (e.g. LAG3) transcripts (4, 8). Notably, groups 2 and 4 exhibited elevated cytotoxic T cell percentages even at T3. Cytotoxic CD4+ T cells should rapidly contract post-disappearance of antigens (27), and so their persistence at T3 suggests an ongoing T cell response (or an anomalously slow contraction) 2-3 months after infection.

**Fig. 2.**
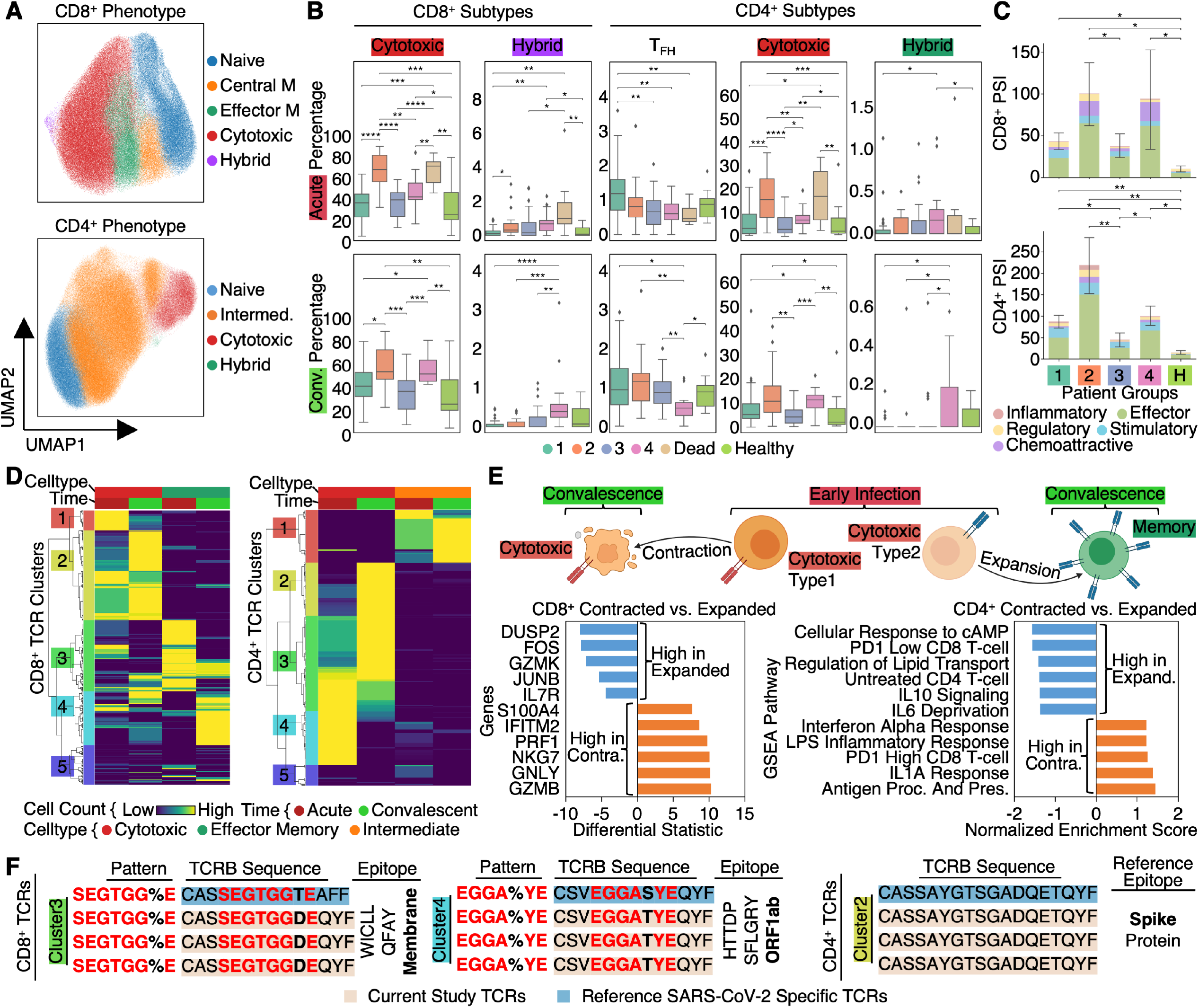
Phenotypical, functional and TCR analysis of CD8+ and CD4+ T cells in patients from the four recovery trajectories. (A) UMAP embedding of CD8+ (top) and CD4+ T cells (bottom) colored by unsupervised clustering. (B) Box plots showing percentages of CD8+ and CD4+ T cell phenotypes in each patient group at acute (T2) (top) and convalescent (T3) (bottom). (C) Single-cell polyfunctional strength index (PSI) of CD8+ (top) and CD4+ (bottom) T cells in each patient group, plotted as mean±SEM. (D) Hierarchical clustering of CD8+ (left) and CD4+ (right) T cell TCRs (rows) based on TCR sharing patterns across select phenotypes and time points (see color key at bottom). Only cytotoxic, effector-memory or intermediate phenotypes are shown. The full heatmap is provided in Fig. S4A. (E) TCR-transcriptome lineage tracking analysis for CD8+ (CD4+) T cells. Top, cartoon illustration for analyzing differential gene signatures for cytotoxic T cells at T2 that will convert into memory (expanded) at T3 versus those will clonally contract at T3. Bottom left, top genes differentially expressed between the two subsets of cytotoxic CD8+ T cells in T2. Bottom right, top pathways differentially regulated at T2 for cytotoxic CD4+ T cells that clonally contracted versus clonally expanded. (F) Representative GLIPH2 groups of TCRs from TCR clusters 3 and 4 (defined in (D), left) for CD8+ TCRs (left two panels) and cluster 2 (defined in (D), right) for CD4+ TCRs (right panel). Blue represents published SARS-CoV-2-specific TCRs, tan represents TCRs from our dataset. Inferred epitopes or virus proteins based on the blue TCR are provided on the right. Boxplot center, median; box limits, 25th and 75th percentiles; whiskers, 1.5× interquartile range (IQR). *p-value<0.05, **p-value<0.01, ***p-value<0.001, ****p-value<0.0001.

Patient group 4 exhibited elevated fractions of the proliferative-exhausted phenotypes at T3 (Fig. 2B). Interestingly, group 4 also showed reduced cytotoxic CD4+ T cell fractions during acute infection (T2) relative to group 2 (Fig. 2B). Continued T cell proliferation and compromised early (T2) T cell responses suggests potential delayed antigen clearance. Group 4 also exhibited repressed levels of T follicular helper (TFH) cells at T3 (Fig. 2B). T¬FH cells facilitate somatic hypermutation and isotype switching in B cells to promote humoral immunity. We return to this issue in our B cell analysis below. Group 2 is distinguished by upregulated Th17 and Treg cells at T3 (Fig. S3G). Upregulated Th17 cells can be linked to autoimmunity risks (28) and upregulated Treg counts, especially activated Treg cells (Fig. S3G), may be controlling the persistent elevation of cytotoxic T cells in group 2 at T3.

### Convalescent patients exhibit higher T cell polyfunctionality than unexposed healthy controls

To evaluate the responsiveness of T cells in recovered patients, we used single-cell secretome technology (29–31) to measure 32 secreted proteins from CD8+ and CD4+ T cells following stimulation. A polyfunctional strength index (PSI) was calculated, reflecting the product of the numbers of different proteins secreted and the secretion copy numbers. Relative to healthy donor T cells, the PSI at T3 was elevated in groups 1, 2 and 4 for both CD8+ and CD4+ T cells (Fig. 2C). This high PSI suggests that T cells at convalescence are more primed to secrete multiple cytokines during a possible recall response. Many of those cytokines possess effector (cytotoxic) functions (Fig. S2H, S3H), but other functional signatures were also detected. As examples, CD4+ T cells from groups 2 and 4 secreted relatively high levels of Th1-like cytokines (e.g. IFN-γ), while CD4+ T cells from group 2 patients uniquely secreted elevated levels of Th2-(IL-4), and Th17-(IL-17A) like cytokines (Fig. S3H). Simultaneous elevation of Th1, Th2 and Th17 within group 2 is consistent with the sc-CITE-seq and suggests a more diverse CD4+ T cell response upon stimulation.

### Clonal and cell-state-transition dynamics reveal divergent groups of TCRs and an early-transcriptome signature associated with memory formation versus clonal contraction

TCR gene sequences provide molecular barcodes to track clonal frequency and transcriptomic alteration from early infection to convalescence. We identified that clonally dominant TCRs at T3 are different from those dominant at T2 (Fig. 2D). We investigated this by separately clustering CD8+ and CD4+ T cell TCRs via their clonal frequencies across distinct transcriptomic phenotypes and time (Fig. 2D, Fig. S4A, table S8, Supplemental Methods). For CD8+ T cells, TCR clusters 3 and 4 were enriched within the effector-memory pool but cluster 3 TCRs were dominant during early infection (T2) and contracted in convalescence (T3), whereas cluster 4 TCRs were clonally dominant at T3 but not T2, for all patient groups (Fig. S4B). Both clusters 3 and 4 contain likely SARS-CoV-2-specific TCRs as revealed by GLIPH2 analysis with published SARS-CoV-2-specific TCRs (32, 33) (Fig. 2F). Certain SARS-CoV-2 epitope specificities are shared across both TCR clusters, while others are cluster specific (Fig. S4C, table S9). Similarly divergent TCR clusters were also revealed in CD4+ T cells (Fig. 2D, F, Fig. S4C).

Using TCRs as lineage-tracing barcodes, we mined for early transcriptomic signatures to predict these TCR dynamics and subsequent cell fates. Specifically, within cytotoxic CD8+ T cells in T2, we queried for transcriptomic differences associated with clonotypes undergoing conversion to memory T cells at T3 versus clonal contraction at T3 (Fig. 2E). “Memory-precursor” cytotoxic T cells at T2 showed biased activation of memory-like genes (e.g. IL7R(34), GZMK(35)), as well as genes that inhibit inflammation or prevent T cell over-activation (e.g. DUSP239, JUNB40) (Fig. 2E bottom-left blue-bars). By contrast, T2 cytotoxic T cells destined for clonal contraction had upregulated genes associated with effector functions (e.g. GZMB, PRF1) and inflammatory-responses (Fig. 2E bottom-left orange-bars). Thus, cytotoxic CD8+ T cells in early-stage COVID-19 with hyper-activation signatures may eventually contract, while subpopulations with early memory-biased signatures will become dominant in the memory pool later. However even at T3, the cytotoxic pool was replenished with new clones (Fig. 2D cluster 2). Similarly, for CD4+ T cells, cytotoxic CD4+ T cells at T2 that upregulate inflammatory-response pathways (e.g. LPS stimulatory response) tended to clonally contract (Fig. 2E bottom-right orange-bars). CD4+ clonotypes that had more regulatory signal responses (e.g. IL-10 signaling) tended towards clonal expansion at T3 (Fig. 2E bottom-right blue-bars). Such early-transcriptomic signatures may potentially provide markers for identifying memory forming TCRs versus short-lived early-protective TCRs. In addition, this analysis implies that some cytotoxic, SARS-CoV-2 specific CD8+ and CD4+ T cell clonotypes continue to emerge at T3, suggesting that viral-antigen presentation may persist 2-3 months post-infection in some patients.

### Anti-SARS-CoV-2 antibody titers observed in convalescent patients correlated with early infection severity

Anti-SARS-CoV-2 antibody responses were elevated for all convalescent patients compare to unexposed healthy donors. We measured IgG, IgA and IgM antibody titers against spike protein, receptor-binding domain (RBD) of spike, and nucleocapsid protein at T3 (Fig. 3A-C, Fig. S5G). Higher IgG and IgA antibody titers against spike or RBD were observed for groups 2 and 4 (Fig. 3aA-C), consistent with severe infection histories in those groups (38, 39). A pseudo-virus neutralization assay (Fig. 3D) showed that neutralizing antibody levels decreased between T2 and T3 in all groups (Fig. 3D middle), and strongly correlated (r=0.77, p=2.2e-16) with anti-RBD IgG titers (Fig. 3D right), as reported (40, 41).

**Fig. 3.**
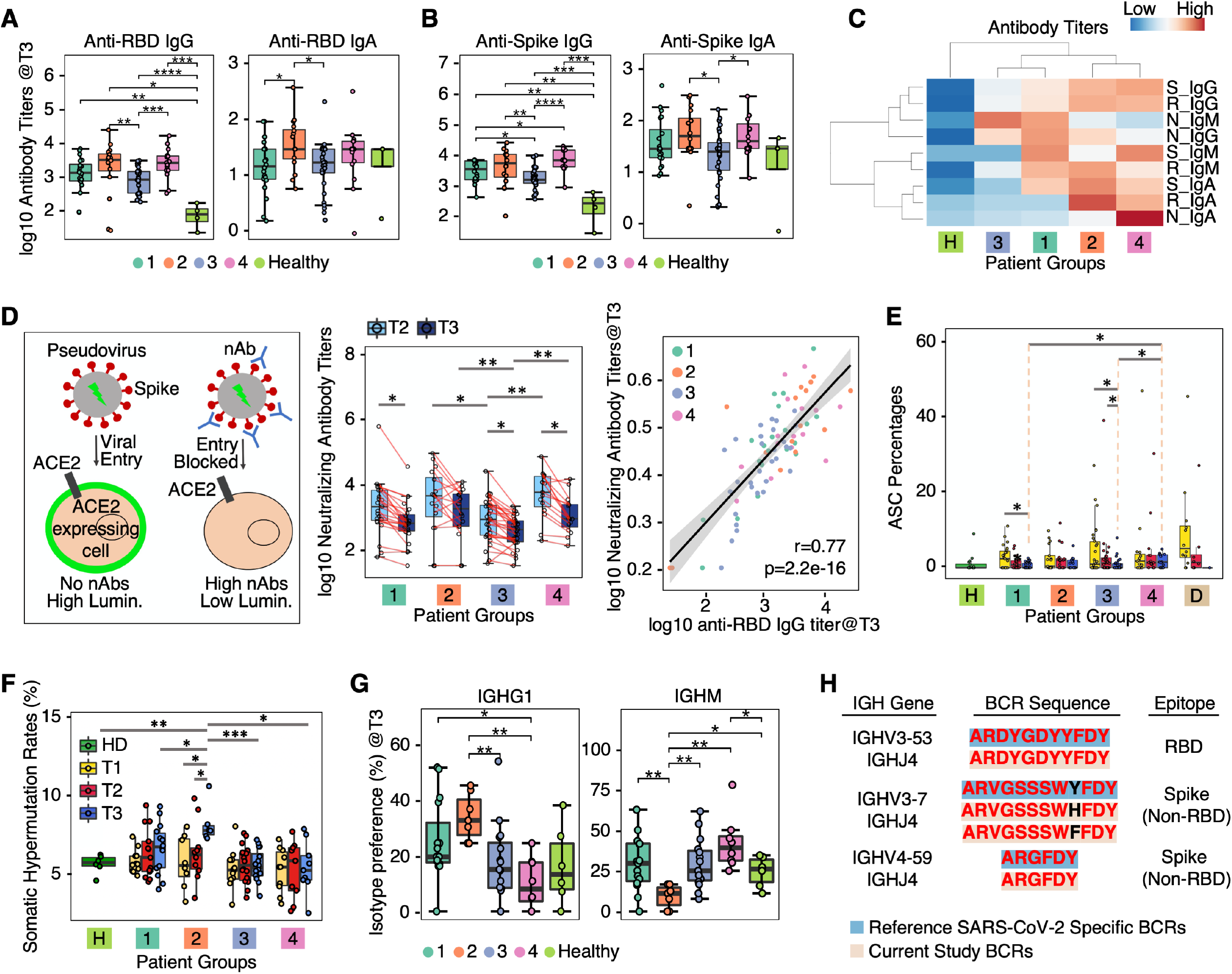
Longitudinal tracking of B cell responses in COVID-19. (A-B), Box plots showing IgG and IgA antibody titers at T3 measured by ELISA to RBD (A) and spike protein (B). (C) Heatmap with hierarchical clustering showing normalized IgG, IgA and IgM antibody titers to RBD, spike and nucleocapsid protein. (D) Pseudovirus neutralization assay of plasma. Left, diagram illustrating the neutralizing assay. Middle, box plot showing neutralizing antibody titers in each patient group at T2 and T3. Samples from the same patient are connected with a red line. Right, linear regression plot with 95% confidence level (grey shaded areas) showing the relationship between neutralizing antibody titers and anti-RBD IgG antibody titers at T3. Pearson correlation coefficient is shown. Points were color-coded by patient group (see color key at top-right). (E) Box plot showing percentages of antibody-secreting cells (ASC) in healthy individuals and each patient group across all time points. (F) Box plot showing somatic hypermutation (SHM) rates in memory B cells across all time points. (G) Box plots showing percentages of IGHG1 (left) and IGHM (right) memory B cells over all memory B cells at T3. (H) Convergent BCR CDR-H3 sequences that are specific for SARS-CoV-2 spike protein shared within some patients in our patient cohort. Boxplot center, median; box limits, 25th and 75th percentiles; whiskers, 1.5× interquartile range (IQR). *p-value<0.05, **p-value<0.01, ***p-value<0.001, ****p-value<0.0001.

### Distinct longitudinal patterns of B cell activation and affinity maturation within patients with severe infection histories

Clustering of B cell single-cell transcriptomes resolved Naïve (IgD+CD20+), intermediate (CD27lowIgDint), memory (CD27+CD20+), and antibody-secreting (ASCs, CD20-CD38+) (Fig. 5SA-E, top) phenotypes. ASCs were elevated in group 4 at T3 (Fig. 3E), indicating prolonged antigen-driven B cell activation and differentiation. Interestingly, proliferative T cells are also higher in group 4 at T3 (Fig. 2B).

Virus-specific memory B cells mature affinity for antigen through accumulation of somatic hypermutations (SHM) (42). For most patients, SHM levels were stable over time, (Fig. 3F) as reported (43). However group 2 exhibited anomalously high overall SHM levels in the memory B cell pool at T3 (Fig. 3F, Fig. S5F), while also showing a preference towards an isotype-switched phenotype (IGHG1) (Fig. 3G). These observations suggest a robust germinal-center (GC)-associated immune response. By contrast, group 4 patients exhibited a high percentage of IGHM memory B cells relative to group 2 (Fig. 3F, G), perhaps suggesting an extrafollicular pathway for B cell differentiation. This can be related to the low percentage of circulating TFH cells in group 4 at T3 (Fig. 2B) (44). However, the ultimate effect on neutralizing antibodies is unclear (Fig. 3D). These distinctions between groups 2 and 4 suggest that factors besides disease severity may influence B cell memory formation.

Of note, we identified convergent BCR sequences with no/low levels of SHM that were reported to be specific for SARS-CoV-2 spike protein (40, 45, 46) in patients at T1 and T2 (Fig. 3H), suggesting that SARS-CoV-2-specific B cells can be generated from naive B cell pool (40, 43, 45, 46).

### Innate immune cell classes in convalescent patients exhibit elevated functionality compared to unexposed healthy donors

We analyzed polyfunctional responsiveness of monocytes and NK cells at T3 following stimulation. We profiled monocytes (Fig. S6A-C) and identified that all recovered patients showed higher levels of polyfunctionality than healthy donors (Fig. S6D, E). For NK cells, group 2 patients showed the highest effector cytokine polyfunctional responses (Fig. S7E, F). This may be associated with elevated percentages of memory NK cells (KLRC2highGZMHhighFCER1GlowPLZFlow) in this group at convalescence (Fig. S7A-D). Elevated functional responsiveness of innate cells in convalescent patients suggests a memory-like behavior of innate immunity, but its potential protective role warrants further investigation.

### Cross-dataset correlations resolve an orchestrated immune response throughout acute infection to convalescence

To provide an integrated view of the distinct immunological trajectories taken by different patient groups, we analyzed pairwise correlations across all metrics; including functional humoral immunity data, clinical measurements, sc-CITE-seq, single-cell secretome data, and plasma-omic data sets across T2 and T3. Hierarchical clustering of these measurements yielded 14 sets, each defined by similar cross-correlation patterns (Fig. 4A). For these 14 sets, similar functions (e.g. polyfunctional responses of monocytes) tended to cluster together into the same set (Fig. 4A right), suggesting coordination within specific immune cell classes. Relative levels of metrics across patient groups are summarized as a heatmap (Fig. 4A top)

**Fig. 4.**
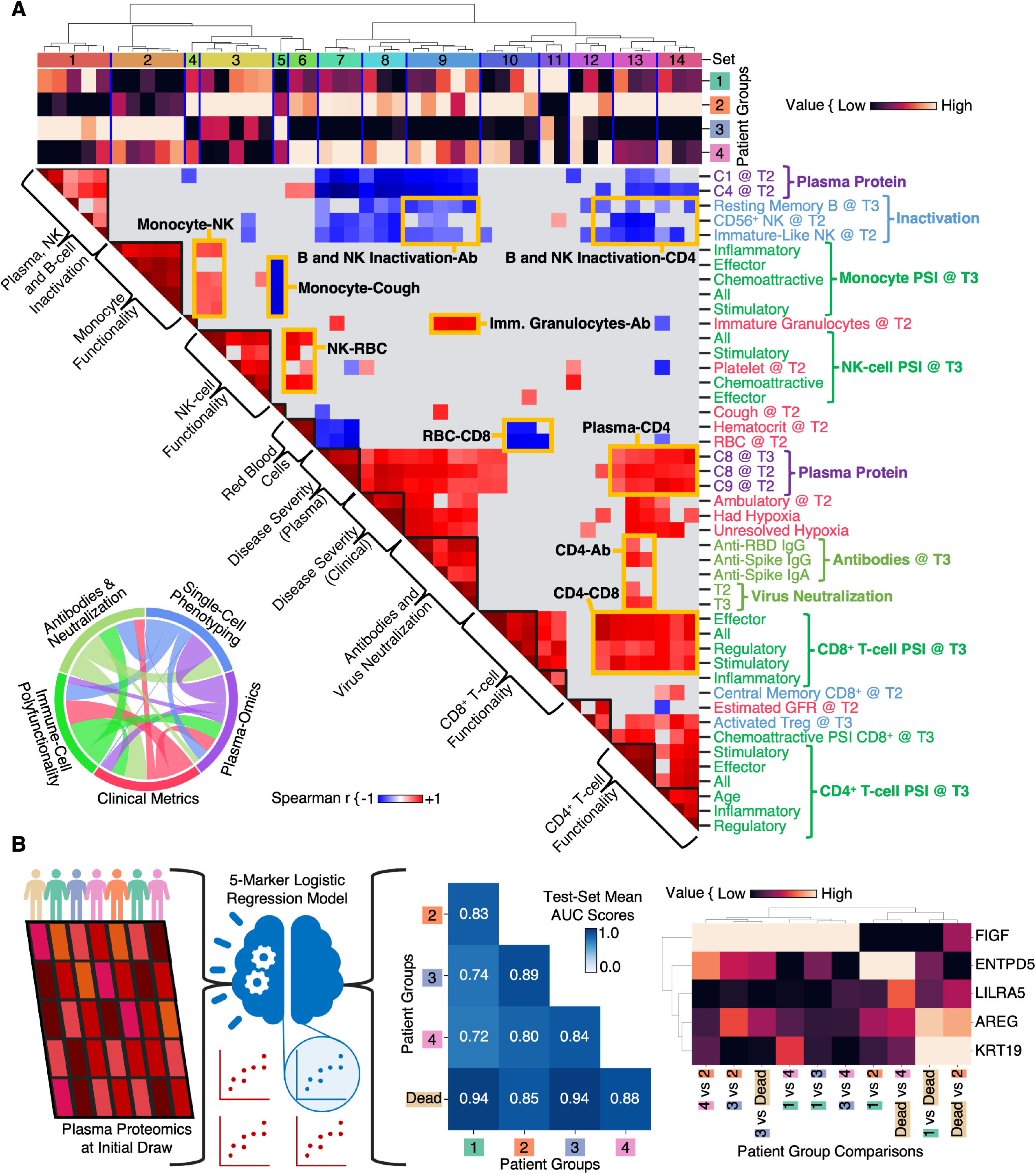
Integrated analysis of functional, phenotypical, clinical data across patients and machine learning model for predicting patient groups at diagnosis. (A) Correlation matrix of 45 functional, phenotypical, plasma and clinical features from COVID-19 patients. Color indicates the Spearman correlation coefficient between row and column variables. Average levels of each measurement for each patient group are shown as the top heatmap. Datasets used for generating this correlation matrix are listed in table S10. (B) Machine learning model for predicting patient groups at diagnosis using five biomarkers. Left, cartoon illustration of machine learning model. Middle, classification accuracy evaluated by AUC scores for pair-wise classification between patient groups using five markers measured at diagnosis. Right, heatmap depicting feature importance of each marker in pair-wise comparisons.

The matrix reveals interesting correlation patterns. Several measures of monocyte PSI at T3 are uniquely high in group 3, and are anti-correlated with whether patients exhibited a cough at T2. CD4+ T cell functionalities (Set 13, 14) exhibited strong correlations with CD8+ T cell PSI metrics (Set 10, 11) and various antibody titers (Set 9) (Fig. 4A, Fig. S8), likely reflecting the key role of CD4+ T cells in orchestrating different arms of immunity. Thus, this correlation matrix illustrates how different immune components coordinate within specific patient groups and further connects that coordination to specific clinical observations.

Further inspection also revealed that levels of plasma protein clusters (C1, C4, C8 and C9) appear to discriminate between patient groups and have extensive correlations with T cell responsiveness, antibody titers and clinical measures (Fig. 4A). This prompted us to identify plasma protein markers to predict immunological recovery trajectory commitment.

### Machine learning model resolves five plasma protein signature at disease diagnosis that accurately predicts patient immunological trajectory

We utilized machine learning to explore plasma proteins that, when measured at diagnosis (T1), were predictive of patient survival from COVID-19 as well as which of the four distinct immunological trajectories the patient would follow (Fig. 4B). We resolved a logistic-regression model that utilized measured levels of five proteins at T1 for predicting all five different outcomes (Supplemental Methods). Pairwise comparisons between outcomes yielded accurate predictions with mean area-under-curve (AUC) values of 0.72-0.94 for validation sets (Fig. 4B middle, Fig. S9A). Each protein measurement is weighted differently per pairwise comparison (Fig. 4B right). The ability of this five-protein panel to resolve all five immunological outcomes suggests that patient trajectories can be predicted at initial clinical diagnosis and underscores the importance of connecting long-term immunological outcomes with early-stage infection status.

## Discussion

This comprehensive profiling of blood plasma and peripheral immune cells of 140 COVID-19 patients is shown as fundamental for categorizing and predicting heterogeneous immunological trajectories during post-disease recovery of COVID-19 (Fig. S10).

Unsupervised clustering of single-cell data revealed four different paths towards recovery, two of which associate with patients who experienced mild COVID-19 (groups 1 and 3), while groups 2 and 4 associate with severe infection histories. Groups 1, 2 and 4 exhibit elevated levels of proteins at T3 that correlate with clinical measures of kidney stress, liver damage and clotting problems (Fig. 1F, G, Fig. S1D). Only patient group 3, representing 36% of our cohort, followed a path of potential return to a healthy baseline at T3. Notably, these patient groups were not distinguished by pre-existing chronic conditions or medications, but could partially be defined by a combination of blood oxygenation level at early-stage disease and patient age (Fig. 1C).

Groups are further distinguished by adaptive and innate immunity. Group 4 exhibited decreased isotype switching and SHM within memory B cells and low levels of TFH cells, pointing to an extrafollicular B cell maturation process potentially resulting from disruption of germinal center formation (47). The differences between group 2 and 4 imply that severe infections don’t guarantee high-affinity class-switched memory B cells, and may suggest the necessity for vaccination of a defined subset of previously infected patients.

Groups 2 and 4 also had elevated fractions of cytotoxic CD8+ and CD4+ T cells at T3 (Fig. 2B), and high T cell polyfunctionality with secreted proteins skewed towards effector molecules (Fig. 2C, Fig. S2G, H, S3H). For both groups, many cytotoxic T cells at T3 were found to exhibit SARS-CoV-2-specific TCR sequences (Fig. 2F). Group 4 patients also exhibited high levels of proliferating T cells. The persistence of cytotoxic and proliferating T cells at T3 may suggest continued exposure to viral antigens. Persistent cytotoxic CD4+ T cells may point to potential immune-pathological risks since they can kill antigen presenting cells (48).

Elevated responsiveness of innate immune cells in recovered patients, together with the elevation of memory-like NK cells in group 2 at T3, implies “memory-like innate immunity” may exist in some patients, but its role of protections from future infections is worth further investigation.

How antiviral T cell populations give rise to T cell subsets with short- and long-lived cell fates is still a fundamental puzzle of T cell immunity (49, 50). Surprisingly, we find that the expanded T cell clones at T2 are different from those at T3. Using TCRs to lineage-trace clonotypes, we found that clones that contract by T3 had previously exhibited elevated markers of over-activation, while T3-expanded clones express genes that inhibit inflammation or prevent T cell over-activation. Whether these clonal dynamics represent more general T cell responses to infection in humans is an interesting question for future study. Further, the fact that new cytotoxic clonotypes continue to emerge at T3 suggests potential ongoing immune responses and may imply incomplete viral clearance in some patients.

Previous studies have mainly focused on biomarkers predictive of patient survival (51). We find that patient survival, as well as which of the four recovery trajectories a patient might take, can all be resolved as early as initial diagnosis, using only a panel of five plasma proteins. Such a biomarker panel may be useful in clinical settings, although further validation on a larger cohort is needed.

Our analyses provide a comprehensive examination of the diverse immunological recovery trajectories for COVID-19. The connections between post-acute outcomes with early infection raise the possibility that interventions early-on may be beneficial in reducing certain outcomes after acute disease. Our analyses show persistence of immune engagement with SARS-CoV-2 and may provide a framework to understand the phenomenon of “long COVID”. Further research is needed to correlate the patient-described and clinician-confirmed symptoms of “long COVID” with our reported immunological disease trajectories. These data may provide a knowledge base for guiding interventional trials to treat and prevent post-COVID-19 symptoms.

## Data Availability

Due to the potential risk of de-identification of pseudonymized RNA sequencing data the raw data will be available under controlled access in the EGA repository upon journal acceptance. For access to data prior to journal acceptance, please contact the corresponding authors.

## Materials and Methods

### Study subjects, WOS score and EHR data extraction

This study included data from 140 COVID-19 patients (63 males and 77 females) and 333 unexposed healthy controls. All enrolled patients provided written informed consent. De-identified proteomic and metabolomic data from matched healthy controls processed using the shared technical pooled control samples to enable batch-correction were previously collected from individuals enrolled in a wellness program (*52*) (Arivale, Seattle, WA). Healthy control samples for single-cell analyses were obtained from Bloodworks Northwest (Seattle, WA). Detailed information on age, sex, race, ethnicity, and disease history etc. of this patient cohort and healthy controls are listed in table S1. Disease severity was quantified using the WHO Ordinal Scale Score (WOS) (*17*). Clinical data for hospitalized patients were abstracted from electronic health records (EHR). Clinical lab data were extracted from the nearest time point to each blood draw. Procedures for the current study were approved by the Institutional Review Board (IRB) at Providence St. Joseph Health with IRB Study Number [STUDY2020000175] and the Western Institutional Review Board (WIRB) with IRB Study Number 20170658.

### Plasma and PBMC isolation

Plasma and PBMCs were isolated from patient whole blood as previously described (*4*). Briefly, plasma fractions were isolated from patient blood collected in EDTA-coated vacutainer tubes (BD, 366643). The PBMC fraction was isolated, counted, and aliquoted at 2.5 million cells/ml in CryoStor CS-10 freeze media (Biolife Solutions, 210102). The aliquoted EDTA-plasma and PBMCs were frozen at -80°C, transferred into liquid nitrogen and stored until use.

### Single-cell multi-omics assay

Chromium Single Cell Kits (10x Genomics, 1000165) were utilized to analyze the transcriptome, surface protein levels, TCR, and BCR sequences simultaneously from the same cell. Experiments were performed according to the manufacturer’s instructions. Briefly, cryopreserved PBMCs were thawed and incubated with the 1X red blood cell lysis solution (Miltenyi Biotech, 130-094-183) to lyse any remaining red blood cells in the PBMC samples. Cells were stained with cell hashtag antibodies (BioLegend, 394661, 394663, 394665, 394667, 394669, 394671, 394673, 394675, 394677, 394679) and TotalSeq-C custom human antibodies (BioLegend, 99814). Stained cells were then loaded onto a Chromium Next GEM chip G (10X Genomics, 1000120). Cells were lysed for reverse transcription and complementary DNA (cDNA) amplification in the Chromium Controller (10X Genomics). The polyadenylated transcripts were reverse-transcribed inside each gel bead-in-emulsion afterward. Full-length cDNA along with cell barcode identifiers were PCR-amplified and sequencing libraries were prepared and normalized. The constructed library was sequenced on the Novaseq platform (Illumina).

### Viral load measurements

The miRNeasy kit (Qiagen) was used to isolate RNA from 100 µl of plasma or nasopharyngeal swab samples according to the manufacturer’s instructions. The RNA was eluted from the membrane with either 30 µl or 50 µl of RNAse free water for plasma or nasopharyngeal swab samples respectively. To detect viral sequences, protocol from the CDC was followed (*53*), and primers were obtained from Integrated DNA Technologies (IDT). The qRT-PCR results were performed on a CFX-96 qPCR machine (Bio-Rad). Levels of SARS-CoV-2 RNA and human RNase P transcript were expressed as cycle threshold (Ct) value.

### Plasma proteomics and metabolomics

Plasma concentrations of proteins and metabolites were measured as previously described (*4*). Batch-corrected proteomic and metabolomic data were further adjusted for age, sex and BMI, as well as their interactions, using a set of robust linear regression models estimated for each protein and metabolite separately using the external control sample of uninfected individuals that were selected using propensity score matching on a number of sociodemographic and comorbidity variables from a larger in-house sample. Models were fitted using the lmrob function from the R package robustbase with the ‘KS2014’ setting (*54*). Metabolite values were log2 transformed prior to further analyses, while protein abundance values (NPX) were already log2 scaled.

### Single-cell multiplex secretome assay

Cryopreserved PBMCs were thawed and incubated in complete medium (RPMI 1640 (Gibco, 11875-093) containing 10% fetal bovine serum (FBS, Gibco, 26140-079), 1x of glutamax (Thermo, 35050061) and 100U/mL penicillin-streptomycin (Gibco, 15140-122)) overnight at 37°C, 5% CO_2_. After overnight recovery, CD4^+^ and CD8^+^ T cells were isolated using CD4^+^ (Miltenyi Biotec, 130-045-101) and CD8^+^ (Miltenyi Biotec, 130-045-201) microbeads sequentially. NK cells and Monocytes were isolated using CD56 MicroBeads, human (Miltenyi Biotec, 130-050-401) and the Pan Monocyte Isolation Kit (Miltenyi Biotec, 130-096-537), respectively.

The isolated CD4^+^ and CD8^+^ T cells were seeded at a density of 1⨯10^5^ cells/well in a 96 well-plate and stimulated for six hours with plate-bound anti-CD3 antibodies (eBioscience, 16-0037-85, pre-coated at 10 µg/ml overnight at 4°C) and 5 µg/mL of soluble anti-CD28 antibodies (eBioscience, 16-0289-85,) in complete medium at 37°C, 5% CO_2_. The isolated NK cells were cultured for 12 hours in the presence of IL-2 (Biolegend, 589104, 10 ng/ml). The enriched monocytes at were seeded at 1⨯10^5^ cells/mL and stimulated with 10 ng/ml lipopolysaccharide (Sigma Aldrich, L2654) for 12 hours. After stimulation, the activated cells were collected, washed, and stained with membrane stain (included in the IsoPlexis kit), before being loaded onto the chip consisting of 12,000 chambers pre-coated with an array of 32 cytokine capture antibodies. The NK cells were resuspended in complete RPMI supplemented with PMA (Sigma, P8139, 5 ng/ml) and Ionomycin (Sigma, 10634, 500 ng/ml) and then loaded onto the IsoCode chip for the stimulation during the incubation. The chip was inserted into IsoLight for further incubation for 16 hours. Secreted cytokines were detected by a cocktail of detection antibodies followed by the fluorescent labeling. Fluorescent signals were analyzed by the IsoSpeak software to calculate the numbers of cytokine-secreting cells, the intensity level of cytokines, and polyfunctional strength index (PSI). Measured cytokines in each panel are listed as below.

Single-Cell Adaptive Immune cytokine panel including the following subsets of cytokines. Effector: Granzyme B, IFN-γ, MIP-1α, Perforin, TNF-α, TNF-β; Stimulatory: GM-CSF, IL-2, IL-5, IL-7, IL-8, IL-9, IL-12, IL-15, IL-21; Chemoattractive: CCL11, IP-10, MIP-1β, RANTES; Regulatory: IL-4, IL-10, IL-13, IL-22, TGFβ1, sCD137, sCD40L; Inflammatory: IL-1β, IL-6, IL-17A, IL-17F, MCP-1, MCP-4.

Single-Cell Innate Immune cytokine panel including the following subsets of cytokines. Effector: IFN-γ, MIP-1α, TNF-α, TNF-β; Stimulatory: GM-CSF, IL-8, IL-9, IL-15, IL-18, TGF-α, IL-5; Chemoattractive: CCL11, IP-10, MIP-1β, RANTES, BCA-1; Regulatory: IL-10, IL-13, IL-22, sCD40L; Inflammatory: IL-1β, IL-6, IL-12-p40, IL-12, IL-17A, IL-17F, MCP-1, MCP-4, MIF; Growth Factors: EGF, PDGF-BB, VEGF.

### ELISAs

Briefly, 384-well plates (ThermoFisher, 464718) were coated with 10 µL of 5 µg/mL SARS-CoV-2 RBD (Invitrogen, RP-87678), spike (S) (Invitrogen, RP-87680), or nucleocapspid (N) (Invitrogen, RP-87707) protein in carbonate pH9.6 buffer overnight at 4°C. Plates were washed four times with wash buffer (phosphate buffered saline (PBS) containing 0.05% Tween-20) and blocked with blocking buffer (wash buffer with 5% BSA) for 1 hour at room temperature (RT). Wells were incubated with 30 µL heat-inactivated plasma samples from COVID-19 patients at six serial three-fold dilutions, starting from 1:30 in blocking buffer for 1 hour at RT. The anti-S antibody (abcam, ab273073) anti-N antibody (abcam, ab272852) at nine serial three-fold dilutions, starting from 2 µg/mL were used as positive controls. A non-coating well, a non-binding well, and a blank well as negative controls wells were also included on the plate. After washing four times with wash buffer, wells were incubated with peroxidase-conjugated goat anti-human IgG (Sigma, A6029, 1:1,000 dilution), IgA (Sigma, A0295, 1:5000 dilution), or IgM (Sigma, A6907, 1:1000 dilution) antibodies in blocking buffer for 1 hour at RT. Wells were washed four times again before incubating with 30 µL 3,3’,5,5’-tetramethylbenzidine (TMB) substrate solution (Seracare, 5120-0047). The TMB reaction was stopped after 5 minutes by adding 1M sulfuric acid. The OD at 450nm was measured on a Spectramax Plate Reader. The ELISA antibody titers were defined as the plasma dilutions that result in the middle response of the positive control and calculated by fitting the background-subtracted data to a four-parameter logistic regression model using the R package nplr (*55*).

### Neutralization assay

The pseudo-virus neutralization assay was conducted by Monogram Biosciences as previously described (*56*). Briefly, pseudo-typed SARS-CoV-2 virus expressing spike proteins was generated based the original Wuhan-Hu-1 strain sequences (GenBank: NC_045512.2). Neutralizing antibody titers were measured by incubating nine serial three-fold dilutions of plasma samples with a starting dilution of 1:40 and SARS-CoV-2 pseudo-typed virus at 37°C for 1 hour. HEK-293 cells expressing ACE2 were added to the 96-well plate and incubated for additional 60-80 hours at 37°C for luminescence measurements. Neutralization titers were calculated as the plasma dilution conferring 50% inhibition (ID50) of pseudo-virus infection, adjusting for background luminescence measured from the SARS-CoV-2 nAb positive control.

### Single-cell sequencing data processing

Droplet-based sequencing data were aligned and quantified via Cell Ranger Single-Cell Software Suite (v3.0.0, 10x Genomics) using GRCh38 as a reference. Cells from each demultiplexed sample were first filtered for cells with ≥200 genes, then filtered based on 1) <10000 unique molecular identifiers (UMI) counts per cell (library size); 2) <2500 detected genes per cell; and 3) proportion of mitochondrial gene counts (mitochondrial gene UMIs / total UMIs)<10%. Doublets were simultaneously identified in sample demultiplexing or using scrublet (*57*) and removed prior to the aforementioned filtering. After QC-based filtering, a total of 846,413 cells were retained for downstream analysis. Scanpy (*58*) was used to normalize cells via CPM normalization (UMI count per cell was set to 10^6^) and log1p transformation (natural log of CPM plus one).

### Single-cell RNA-seq cell type identification

Normalized, ln(CPM+1), mRNA data from QC-passing single cells were analyzed via PCA (ARPACK). All 50 PCs were used to calculate a neighborhood graph (n_neighbors=15) which was utilized to determine UMAP (*59*) coordinates and Leiden clusters (*60*). Clusters were assigned cell types based on canonical immune markers and multi-cell-type clusters were separated via additional UMAP and Leiden cluster calculations. Clusters (14,251 cells) that co-expressed markers from multiple cell types were labeled as low-quality or doublets and removed from further analysis. In total, 832,162 cells were deemed high-quality and assigned cell types; these cells did not show noticeable batch differences.

Labeled T cells were used to calculate a CD4^+^ score (sum of min-max-scaled normalized levels of *CD4* transcript and CD4 surface protein) and a CD8^+^ T cell score (sum of min-max-scaled normalized levels of *CD8A* and *CD8B* transcripts, and CD8 surface protein). The two scores were min-max-scaled and then projected for manual gating of CD4^+^ and CD8^+^ T cells. T cells with ambiguous scores were classified as “Other T cells”.

### Single-cell regulon-based phenotype identification

Normalized mRNA values for each major immune cell types (B cells, CD4^+^ T cells, CD8^+^ T cells, monocytes and NK cells) were used to construct regulon values via pySCENIC (*61*). Patient 4 B cells were removed pre-pySCENIC-calculation as they had a cancerous (chronic lymphocytic leukemia) population of B cells. Single cell regulon matrices per cell type were utilized to calculate PCA values (50 PCs). PCs that represent technical factors determined via correlation with n-counts per cell or sequencing batches were removed; specifically, PC1 for B, CD4, monocyte and NK populations and PC5 for CD8. Remaining PCs were used for neighborhood graph (n_neighbors=15) then UMAP and Leiden cluster calculations with resolution input of 0.9, 0.85, 1.2, 0.95 and 1.0 for B, CD4, CD8, monocyte and NK, respectively. Cells were then additionally screened for potential doublets; low-quality cells were only detected in B cells where Leiden clusters 7 and 9 were deleted then dimensional reduction and clustering were re-done with PC1 removed due to technical correlation and a Leiden resolution of 0.7. A separate clustering on just memory B cells identified from previous clustering was done to further separate B cell classes using a Leiden resolution of 0.5.

CD4^+^ T cells were assigned phenotypes based on canonical marker annotated Leiden clusters with Naïve (Leiden 0); Intermediate (2,3,5,6); Cytotoxic (4) and Hybrid (7). Additional phenotypes T_FH_, Treg and Th_17_ were assigned if cells contained normalized mRNA levels above 2.8 (determined via bimodal distribution of mRNA levels from a density plot) for *CXCR5, FOXP3*, or *RORC*, respectively, and were not already assigned as a Cytotoxic or Hybrid cell. CD8^+^ T cells were similarly labeled with canonical marker described Leiden clusters with Naïve (Leiden 0,10,11,12); Central Memory (6); Effector Memory (4,8); Cytotoxic (1,2,3,5,7,9,13) and Hybrid (14). B cells were annotated as Naïve B cells (Leiden 0), memory B cells (1), intermediate Naïve and memory B cells (2), and plasma cells (4). Memory B cells were further annotated with atypical memory B cells (Leiden 2), resting memory B cells (0), and activated memory B cells (1). Monocyte and NK Leiden clusters were annotated based on differential gene expression (Wilcoxon test) and canonical markers. All reduced dimensions (PCA, neighborhood graph, UMAP) and clusters (Leiden) for all of the single cell RNA-seq data were calculated via Scanpy (*58*).

### Hierarchical clustering of patient trajectories

Transcription factor module values of the single cells for the aforementioned immune classes B, CD4, CD8, monocyte, and NK cells were classified into nine Leiden clusters via a resolution input for Leiden clustering of 0.75, 1.0, 1.0, 0.95 and 1.0, respectively. Percentages of Leiden clusters were calculated per patient timepoint (i.e. blood draw) for each cell type. T2 (acute) and T3 (convalescent) percentages of cell type Leiden clusters were extracted and utilized to cluster patients via Ward’s method with criterion “maxclust” and t=4, as visually determined.

### Decision tree analysis

Classification and Regression Trees (CART) recursive partitioning was used to construct a decision tree for identifying patient groups. Independent variables included age, sex, chronic conditions (hypertension, chronic kidney disease, heart failure, chronic kidney disease, diabetes), count of those chronic conditions, medications (immunosuppressants, angiotensin converting enzyme inhibitors, and angiotensin II receptor blockers), minimum blood oxygenation, dyspnea, limitations on activity and disease severity (maximum WOS). For the given set of 15 clinical factors, there were no major observable difference between group 2 and 4 and they were thus combined. Hence, for the CART analysis three patient groups were used as dependent variables (1, 3, 2 and 4). Ten-fold cross-validation was applied to evaluate tree reliability, and gini index was used to quantify the impurity and select splits during classification. To avoid overfitting, a maximum acceptable difference in risk between the pruned and the sub-tree was set at one standard error. Missing data were handled by surrogate splits. The following formula were used to calculate misclassification rate or prediction error in cross-validation (root node error x relative error x 100) and prediction accuracy (1 – missclassification rate).

### Plasma proteomic Gene Ontology (GO) analysis

Batch-corrected plasma protein levels were converted into Z-scores using the means and the standard deviations estimated for the residuals in the matched control samples, which included corrections for age, sex, and body mass index (BMI). The Z-scores thereby indicate the difference in plasma abundance relative to the control sample - e.g., a Z-score of 2 represents 2 standard deviations above the expected control value. The Z-scores were used for plotting the line plot in Fig. 1e middle panel. A score for a GO process was calculated by first summing the averaged Z-scores from GO-affiliated plasma proteins with a Z score larger than 2 or smaller than -2. Those proteins were termed significant outliers. The resulting sum was multiplied by the fraction of significant outliers relative to all possible measured proteins associated with that GO process to yield a GO score. For example, for Developmental Processes, Group 4 patients, T3 time point: 64 proteins are positive outliers, and one protein is a negative outlier, with Z-scores summing to 192.9. There were 243 proteins measured that were associated with this GO process, so 192.9×65/243=GO Score of 51.6. Such scores were used for plotting Fig. 1d bottom panel.

### Clinical factor and plasma analyte correlation analysis

We estimated Pearson correlations between clinical information and EHR data (clinical factors) and plasma abundance of proteins and metabolites. Clinical information and lab tests were extracted from EHRs; continuous variables were directly utilized, and categorical variables were binarized when possible. Multi-class categorical variables were binarized by performing separate comparison between each non-standard category with the standard. For example, the multi-class variable “hypoxia” contained categories resolved, unresolved and no hypoxia leading to the comparisons resolved vs. no hypoxia, unresolved vs. no hypoxia, and had hypoxia (unresolved or resolved) vs. no hypoxia. In all cases no hypoxia was set to 0 and the other category was set to 1. All analyte values were from acute (T2) measurements.

Correlations that had |r|≥0.5 (magnitude of r), p-value<0.05 and n-samples>15 (number of blood draws used for correlation) were deemed significant. Remaining x (plasma analyte) and y (clinical factors) were further filtered via their number of significant correlations. Lists of x and y analytes were progressively filtered by removing analytes until all analytes had at least three significant correlations with others. Resulting correlation heatmap was ordered by non-filtered correlation values and heatmap was masked by only showing the significant correlations. Clustering was done via Ward’s method with criterion “maxcluster”, t “8” (“5”) for plasma (clinical factors), as visually determined.

Protein scores were calculated via standard scaling (subtract mean, divide by standard deviation) the proteins within defined plasma analyte clusters then averaging the scaled values per patient per plasma analyte cluster.

### Differential metabolites across patient groups

Metabolites with all NaN values were removed and only values from healthy donors or convalescent draws of patients from patient groups were kept. Differential expression of metabolites was computed via scanpy.tl.rank_genes_groups (method=“Wilcoxon”, n_genes=all variables). Each metabolite was assigned to the group with the highest differential score; the second-highest differential score was subtracted from the highest to give a normalized differential score which ranked metabolites intra-group, the top30 metabolites per group were taken. Intra-group ordering of metabolites were done via the same methods as the previous correlation analysis.

### Inter-omic correlation analysis

Analytes and values included in Spearman correlation analysis are provided in table S10. Significant correlations were defined, and ordering and clustering performed in the same manner as the previous correlation analysis with t =14. The top5 analytes (ranked via n-significant-correlations) from plasma clusters, 10x-derived phenotypes, clinical information, lab tests and top25 analytes from functional data were selected.

### Single-cell TCR-seq data processing

Droplet-based sequencing data for T cell receptor sequences were aligned and quantified using the Cell Ranger Single-Cell Software Suite (version 3.1.0, 10x Genomics) against the GRCh38 human VDJ reference genome.

### Single-cell TCR phenotype associations

Filtered annotated contigs for TCRs were analyzed via scirpy (*62*). Aforementioned contigs were filtered for either CD4^+^ or CD8^+^ T cells (as identified via single cell RNA-seq analysis) and then subject to clonotype definition and clonal expansion analysis utilizing nucleotide sequences. Samples were then concatenated together and merged with gene expression data for simultaneous single cell TCR and RNA data visualization.

Both the integrated CD4^+^ and CD8^+^ T cell datasets were subject to filtering for cells with complete TCR sequences, defined as a detectable TRA and TRB. TCRs were normalized per sample (patient blood draw) by sampling with (without) replacement TCRs of samples with n-TCRs < (≥) median TCRs per sample. Pheno-tags were created by compounding cell phenotype with blood draw timepoint (filtered for acute and convalescent). TCR x pheno-tag matrix was constructed with values as the percent of cells in the given pheno-tag with the given TCR. Only TCRs present in ≥2 pheno-tags were included, and values were normalized to ln(value+1). The matrix was then ordered and clustered in the same manner as the correlation analyses with t set to “5”, as visually ascertained.

### Differential gene expression and pathway enrichment analysis of TCR clonal trajectories

For CD8^+^ T cells, TCR group 1 and 5 were was considered clonally deleted and expanded respectively. Differential analysis was performed via scanpy.tl.rank_genes_groups (method=“Wilcoxon”, n_genes=300) on single cells comparing clonally deleted vs. expanded cells. For CD4^+^ T cells, TCRs were hyper-clustered with t =14 and cluster 5 and 10 were considered clonally deleted and expanded respectively. Pseudo-bulk values per clonal-trajectory (averaged across single cells) were utilized in GSEA analysis to derive pathway scores.

### GLIPH2 Analysis

The TCRs of interest were run with a database to cluster TCRs with TCRs of known specificity. Two distinct GLIPH2 were run. For CD8^+^ T cells, we utilized a database from Adaptive Immunity of TCRs matched to SARS-CoV-2 epitopes (*63*). For CD4^+^ T cells, we utilized a database of CD4^+^ T cells that were reactive to SARS-CoV-2 peptides. In both cases, GLIPH2 was run without a reference. The output was modified to only include clusters that shared a V gene. The output was visualized as a binary heat map where positive=at least 1 TCR from the cluster occurred in a GLIPH2 group with the respective database TCR.

### Single-cell BCR & RNA-seq integration

Annotations from sc-RNAseq were used to define memory B cells in the sc-BCR data. Somatic hypermutation rates (SHM) were defined as the percentages of gaps and mismatches in the query sequence compared to the top germline V gene hit identified through IgBLAST (*64*) for each contig. Filtered contig outputs from the 10x Genomics Cell Ranger pipeline were used as input to the R package Immunarch (*65*) to assign clonotypes to memory B cells for each T3 blood draw for calculation of isotype usage in Fig. 3g.

### BCR convergent sequence analysis

IGH sequences from single cells with paired productive heavy and light chains were searched against a list of known SARS-CoV-2 binding antibodies to identify convergent sequences according to the following criteria: utilization of the same IGHV and IGHJ genes; same CDR-H3 lengths; and CDR-H3 amino acid sequences that were within a Hamming distance cutoff of 15% of the length of the CDR-H3.

### Machine learning

Z-scores of plasma protein abundance at baseline (T1) were used to construct a binary classifier to predict patient group status (including death). Analytes were pre-filtered for Z-score≥2 in any patient group or Z-score-difference≥2 between any patient group pair. For a given random state, the top15 most important features derived from ExtraTreesClassifier (n_estimators=100) per pair were concatenated into a list of top features. Top15 most represented features from aforementioned list was used to derive all possible five marker combinations. Each combination was tested via StratifiedShuffleSplit cross validation (CV) with train,test=2/3,1/3 percentages. For each train,test combination GridSearchCV (scoring=“accuracy”) was utilized with logistic regression and range of C’s from 10^−2^ to 10^13^ using StratifiedShuffleSplit in the same manner as before, AUC scores were recorded via sklearn’s roc_curve and auc methods.

Models (marker combination and random state) were then filtered by the average minimum AUC of pair-wise comparisons across CVs and ranked by the number of CVs that have a minimum AUC > 0.7 resulting in the model with random state 2 using markers FIGF, KRT19, ENTPD5, LILRA5 and AREG. Marker significance was then approximated via its coefficient in the logistic regression model corresponding to a given pair-wise comparison. Mean AUC scores were obtained by averaging AUCs across CV iterations and ROC curves were plotted via sklearn’s plot_roc_curve per CV iteration and binary classification.

### Statistical analysis

Statistical analyses were performed using Python or R software packages. Null hypotheses between two groups used in all bar plots and box plots were tested using the non-parametric Mann-Whitney U test. Other specific statistical tests and their significance levels were denoted in each figure legend.

## Acknowledgments

We appreciate the insightful discussion from Prof. David Baltimore, Prof. David Koelle, Prof. Alan Aderem and the ISB COVID-19 Study Group. We are grateful to all participants in this study and to the medical teams at Swedish Medical Center for their support. We thank the Northwest Genomic Center for help with sequencing services, and the ISB-Swedish COVID-19 Biobanking Unit. We thank Amazon Web Services for their support through cloud computing credits provided by the AWS Diagnostic Development Initiative (DDI). We acknowledge funding support from the Wilke Family Foundation (JRH), the Murdock Trust (JRH), Gilead Sciences (JRH), the Swedish Medical Center Foundation (JDG), the Parker Institute for Cancer Immunotherapy (JRH, MMD, PDG, LLL, AR and JAB), Merck, and the Biomedical Advanced Research and Development Authority (HHSO10201600031C to JRH). KW was funded by DOD (W911NF-17-2-0086), NIH (R01 DA040395 and UG3TR002884). RG was funded by the NIH Human Immunology Project Consortium (U19AI128914) and the Vaccine and Immunology Statistical Center (Bill and Melinda Gates Foundation OPP1032317). Further funding by NIH (AI068129 to LLL and R21 AI138258 to NS). YS was supported by the Mahan Fellowship at Herbold Computational Biology Program of Fred Hutch Cancer Research Center.

## Author contributions

Conceptualization: YS, JDG, JRH; Methodology: YS, DY, DGC, JRH; Formal Analysis: YS, DGC, DY, AJPB, SAK, RG, JRH; Investigation: YS, DY, DGC, JC, CLD, SH, RZ, JX, SL, KS, AJPB, SD, CL, RHN, SAK, PB, VRD, KGA, JL, FY, CR, PT, BS, JZ, SM, KM, RE, LJ, YZ, LR, RL, WC, DSOM, CRD, JAW, HAA, WW, NDP, NS, JEH, ATM, KW, AR, LLL, SDB, JAB, LH, RG, PDG, MMD, JDG JRH; Resources: JDG, JRH; Writing – Original Draft: YS, DY, DGC, JRH; Writing – Review & Editing: YS, DY, DGC, JC, CLD, SH, RZ, JX, SL, KS, AJPB, SD, CL, RHN, SAK, PB, VRD, KGA, JL, FY, CR, PT, BS, JZ, SM, KM, RE, LJ, YZ, LR, RL, WC, DSOM, CRD, JAW, HAA, WW, NDP, NS, JEH, ATM, KW, AR, LLL, SDB, JAB, LH, RG, PDG, MMD, JDG, JRH.

---

**Fig. S1. Overview of the Longitudinal Multi-Omic Analysis of Immune Responses in COVID-19 Patients**

**(A)** Cartoon illustration of patient recovery trajectory calculation based on immune cell subpopulation percentages from early infection (T2) to recovery (T3).

**(B)** Bar plot showing age, viral load or clinical metrics across patient groups at acute (T2) or convalescence (T3). P-values were FDR-corrected.

**(C)** Analysis of group-unique metabolites at T3 for patient groups and healthy unexposed individuals. Top, heatmap of top30 metabolites uniquely upregulated per group. Columns indicate metabolites and rows indicate patient groups at T3 or unexposed healthy donors (see color key at bottom). Bottom, box plots showing plasma abundance of selected metabolites per group and levels across four patient groups at T3 or of healthy donors.

**(D)** Inter-omic correlations between plasma-omic analytes and clinical data (full version of Fig. 1f). Left, heatmap of cross-omic correlations. Each row (column) corresponds to a clinical measurement from electronic health records (plasma protein or metabolite). Color represents Pearson correlation coefficient of row and column variable (see color key at bottom).

Boxplot center, median; box limits, 25^th^ and 75^th^ percentiles; whiskers, 1.5× interquartile range (IQR). Right, abundance of plasma analyte cluster across patient groups at T3. *p<0.05, **p<0.01, ***p<0.001, ****p<0.0001.

**Fig. S2. Phenotypical and functional analysis of CD8**+ **T cells in patients with four different recovery trajectories**.

**(A-C)** UMAP embedding of CD8^+^ T cells colored by unsupervised clustering (left panel in **A**), levels of selected mRNA transcripts (other panels in **A**), surface proteins (**B**) and clonal size (**C**).

**(D)** Clonal expansion score for CD8^+^ T cells of samples from patient groups and healthy unexposed individuals at acute (T2) (top) and convalescent (T3) (bottom).

**(E)** UMAP embedding density of CD8^+^ T cells per patient group and time point (T2 and T3). Selected clusters are encircled (see color key in **A**).

**(F)** Box plots showing CD8^+^ T cell phenotype percentages at acute (T2) (top) and convalescent (T3) (bottom) in each patient group.

**(G)** Box plots indicate percentage of CD8^+^ T cells secreting selected cytokine/chemokines for samples from different patient groups at convalescence or from unexposed healthy donors.

**(H)** Heatmap visualization of average cytokine secretion frequency for cells from patient groups at convalescence or healthy unexposed individuals.

Boxplot center, median; box limits, 25^th^ and 75^th^ percentiles; whiskers, 1.5× interquartile range (IQR). Right, abundance of plasma analyte cluster across patient groups at T3. *p<0.05, **p<0.01, ***p<0.001, ****p<0.0001.

**Fig. S3. Phenotypical and functional analysis of CD4**+ **T cells in patients from the four recovery trajectories**.

**(A-C)** UMAP embedding of CD4^+^ T cells colored by unsupervised clustering (left panel of **A**), levels of selected surface proteins (other panels of **A**), mRNA transcripts (**B**), and clonal size (**C**).

**(D)** Clonal expansion score for CD4^+^ T cells of samples from patient groups and healthy unexposed individuals at acute (top) and convalescent (bottom).

**(E)** UMAP embedding density of CD4^+^ T cells for samples across patient groups and time (T2 and T3). Selected clusters are encircled (see color key in **A**).

**(F)** Bar plot of clonal expansion levels per CD4^+^ T cell phenotype color-coded by clonal expansion sizes (n=1, n=2-4, n≥5).

**(G)** Box plots showing the percentages of CD4^+^ T cell phenotypes at acute (T2) (top) and convalescent (T3) (bottom) stages for each patient group.

**(H)** Box plots indicate percentages of CD4^+^ T cells secreting selected cytokine/chemokines for samples across patient groups at convalescence or from healthy unexposed donors.

Boxplot center, median; box limits, 25^th^ and 75^th^ percentiles; whiskers, 1.5× interquartile range (IQR). *p-value<0.05, **p-value<0.01, ***p-value<0.001, ****p-value<0.0001.

**Fig. S4. Lineage tracking revels TCR clusters of distinct clonal dynamics and early signatures for memory T cells formation versus clonal contraction**.

**(A)** Hierarchical clustering of CD8^+^ (left) and CD4^+^ (right) TCRs (rows) based on TCR sharing patterns across phenotypes and time (columns, see color key on right of heatmaps). Full heatmap with all T cell phenotypes for Fig. 2g.

**(B)** TCR clonal shifting patterns from T2 to T3 across patient groups. Box plots showing the percentages of TCRs from each TCR cluster (defined in **A**) over all TCRs for each patient group at acute (T2) and convalescent (T3) time points, for CD8^+^ (left two panels) and CD4^+^ (right two panels) T cells.

**(C)**Hierarchical clustering of inferred SARS-CoV-2 epitopes for each TCR from CD8^+^ (left) and CD4^+^ (right) T cells. Rows, TCR clusters from **A**. Columns, inferred epitopes or viral proteins specific for each TCR (left) or reference SARS-CoV-2 specific CD4^+^ TCR (right). Beige color indicates given TCR cluster contains column’s viral epitope/protein.

Boxplot center, median; box limits, 25^th^ and 75^th^ percentiles; whiskers, 1.5× interquartile range (IQR). *p-value<0.05, **p-value<0.01, ***p-value<0.001, ****p-value<0.0001.

**Fig. S5. B cell responses in COVID-19**

**(A-C)** UMAP embedding of B (top) and memory B cells (bottom) color-coded by unsupervised clustering **A**, levels of mRNA transcripts (**B**) and surface proteins (**C**).

**(D-E)** Heatmap showing normalized expression of selected mRNA transcripts (**D**) and surface proteins

(**E**) per cluster (defined in **A**) in B (top) and memory B cells (bottom).

**(F)** Box plots showing somatic hypermutation rates in Ig gamma heavy chains (left), mu heavy chains (middle) and light chains (right) of memory B cells at T3.

**(G)** Box plots showing IgM antibody titers against RBD, spike and nucleocapsid protein, and IgG and IgA antibody titers against the nucleocapsid protein measured at T3.

Boxplot center, median; box limits, 25^th^ and 75^th^ percentiles; whiskers, 1.5× interquartile range (IQR). *p-value<0.05, **p-value<0.01, ***p-value<0.001, ****p-value<0.0001.

**Fig. S6. Phenotypical and functional analysis of monocytes**.

**(A)** UMAP embedding of monocytes colored by unsupervised clustering (left panel) and levels of selected mRNA transcripts and surface proteins (other panels).

**(B)** UMAP embedding density of monocytes for samples from different patient groups at acute (T2) and convalescent (T3) time points. Selected clusters are encircled (see color key in **A**).

**(C)** Box plots showing the percentage of non-classical monocytes and myeloid-derived suppressor cells (MDSCs) at acute (T2) and convalescent (T3) stages for each patient group. Data are represented as mean±SEM.

**(D)** Single-cell polyfunctional strength index (PSI) of monocytes in each patient group. Data are represented as mean±SEM.

**(E)** Heatmap visualization of average cytokine secretion frequency for monocytes for each patient group at convalescence or healthy unexposed individuals.

*p-value<0.05, **p-value<0.01, ***p-value<0.001, ****p-value<0.0001.

**Fig. S7. Phenotypical and functional analysis of NK cells**.

**(A-B)** UMAP embedding of NK cells colored by unsupervised clustering (left panel of **A**) and levels of selected surface proteins (other panels of **A**) and mRNA transcripts (**B**).

**(C)** UMAP embedding density of NK cells for samples in each patient group and time points (T2 and T3). Selected clusters are encircled (see color key in **a**).

**(D)** Box plots showing the percentages of NK cell phenotypes at acute (T2) and convalescent (T3) stages for each patient group.

**(E)** Bar plot showing *GZMH* transcript levels (left) and NK cell effector PSI (right) in each patient group and healthy unexposed individuals. Data are represented as mean±SEM.

**(F)** Heatmap visualization of average cytokine secretion frequency for NK cells for each patient group at convalescence or healthy unexposed individuals.

Boxplot center, median; box limits, 25^th^ and 75^th^ percentiles; whiskers, 1.5× interquartile range (IQR). *p-value<0.05, **p-value<0.01, ***p-value<0.001, ****p-value<0.0001.

**Fig. S8. Correlations between different measurements**

Scatter plots depicting correlation among different measurements across patients with 95% confidence intervals shaded in blue. Spearman’s correlation coefficients and p-values are shown. *p-value<0.05, **p-value<0.01, ***p-value<0.001, ****p-value<0.0001.

**Fig. S9. Machine learning model evaluation on validation sets and levels of five biomarkers at T1 across patients**.

**(A)** Receiver operating characteristic curves, per cross-validation (CV) iteration, for pair-wise classification (see subtitles) based on the levels of 5 markers at T1 for different validation pairs. Area-under-curve (AUC) values for different CVs (in different colors) are displayed.

**(B)** Boxplots showing plasma abundance of the five plasma biomarkers levels across the four patient groups or deceased patients at T1.

Boxplot center, median; box limits, 25^th^ and 75^th^ percentiles; whiskers, 1.5× interquartile range (IQR).

**Fig. S10. Summary of immune features across four convalescent patient groups and healthy donor**.

**Table S1. Clinical data for each patient and healthy donor**

Clinical measurements and observations of each blood draw of COVID-19 patients and healthy donors included in our study are shown. The Study Subject IDs in each of the four immunological trajectory groups identified in our study are included as well.

**Table S2. Immune cell subpopulation percentages**

The percentages of each immune subpopulation of B cells, CD4^+^ T cells, CD8^+^ T cells, monocytes and NK cells for each sample are shown.

**Table S3. Patient number in each leaf node**

Numbers of patients of each group in each leaf node in **Fig. 1C** are shown. The group with majority of patients and percentages of the major patient group in that leaf node are shown.

**Table S4. Viral load of plasma and nasal swabs**

Cycle threshold (CT) values of plasma viral load (RNAemia) and nasal swab viral load of each sample are shown. A CT value > 40 is labeled with “ND” and a missing value due to the unavailability of the sample is labeled with “Not available”.

**Table S5. Plasma proteomics and metabolomics data**

The batch-corrected proteomics and metabolomics data adjusted for age, sex, and BMI are included. The proteomics data further converted to Z-scores are also included.

**Table S6. Top 30 upregulated metabolites in each patient group**

The top 30 upregulated metabolites per group calculated through differential expression are shown.

**Table S7. Plasma analyte clusters**

Proteins and metabolites in each of the nine plasma analyte clusters in **Fig. 1F** are shown.

**Table S8. TCR clusters**

The CDR3 nucleotide sequences of paired TCRα and TCRβ chains in each TCR cluster in **Fig. 2D**. CD8 and CD4 TCR Clusters are shown separately.

**Table S9. SARS-COV-2 epitopes identified via GLIPH2 analysis for TCR clusters**

The amino acid sequences of SARS-COV-2 epitopes that the TCR clusters in **Fig. 2D** have a specificity for are shown. A “1” for a cluster indicates a TCR specific for that epitope could be identified in this cluster.

**Table S10. Datasets used for generating the correlation matrix in Fig. 4A**

282 features derived from multiple datasets in our study for each patient are shown. Cell type, assay type, and the time of the blood draw are indicated as well.

